# Fine scale spatial mapping of urban malaria prevalence for microstratification in an urban area of Ghana

**DOI:** 10.1101/2025.03.03.25322260

**Authors:** Samuel Kweku Oppong, David Kwame Dosoo, Nana Yaw Peprah, George Asumah Adu, Wahjib Mohammed, Jen Rozier, Kingsley Kayan, Michael McPhail, Punam Amratia, Kefyalew Addis Alene, Kwaku Poku Asante, Peter W. Gething, Keziah L. Malm

## Abstract

**Background:** Malaria in urban areas is a growing concern in most sub-Saharan African countries. The growing threats of *Anopheles stephensi* and insecticide resistance magnify this concern and hamper elimination efforts. It is therefore imperative to identify areas, within urban settings, of high-risk of malaria to help better target interventions.

**Methods:** In this study, we combined a set of environmental, climatic, and urban covariates with observed data from a malaria prevalence study and used geospatial methods to predict malaria risk in the Greater Accra Region of Ghana. Georeferenced data from 12,371 surveyed children aged between 6 months and 10 years were included in the analysis.

**Results:** Predicted malaria prevalence in this age group ranged from 0 to 52%. Satellite-driven data on tasselled cap brightness, enhanced vegetation index and a combination of urban covariates were predictive of malaria prevalence in the study region. We produced a map that quantified the probability of malaria prevalence exceeding 10%.

**Conclusions:** This map revealed areas within the districts earmarked for malaria elimination that have high malaria risk. This work is providing evidence for use by the National Malaria Elimination Program and District Health Managers in planning and deploying appropriate malaria control strategies.

**Summary box:** *What is already known?:* Reduction in malaria incidence globally has stalled in the past few years. Malaria endemic countries are being encouraged to use local data to inform appropriate malaria control strategies. Malaria prevalence studies seldomly provide estimates below regional administrative levels. The availability of environmental, climatic, and socioeconomic factors as well as computational methods has enhanced predictive methods that quantifies the disproportionate variation of malaria risk between and within urban areas.

*What are the new findings?:* Predictive maps of malaria at high spatial resolutions such as 100m allows for visualizing fine-scale heterogeneity of malaria in neighbourhoods. Inclusion of urban covariates in models predicting malaria risk in urbanized communities helps to account for socioeconomic disparities and their effect on malaria risk.

*What do the new findings imply?:* Malaria control efforts needs to be guided by highly granular data. Systems to generate granular data on a continuous basis needs to be strengthen in malaria endemic countries, especially, to better inform deployment of appropriate interventions in resource constraint settings. This type of analysis provide information on which intervention is appropriate in a specified geographical area.

## Introduction

Malaria remains a critical global health challenge, with an estimated 247 million cases and over 600,000 deaths reported annually, the vast majority in sub-Saharan Africa(1). Although significant strides have been made in reducing the burden of malaria over the past two decades, progress has slowed in many regions. Malaria has historically been considered a rural disease, closely linked to natural water sources and vector habitats(2). However, urban malaria is becoming an increasing concern, particularly in malaria-endemic regions such as Ghana, where rapid urbanization has introduced new challenges for disease control(3). Urbanization is a global phenomenon, with over half of the world’s population now residing in urban areas, a figure that is expected to rise to 68% by 2050(4,5). In sub-Saharan Africa, the urban population has surged from 39% in 2003 to 53% in 2021(6). This rapid, and often unplanned, urbanization has led to the proliferation of informal settlements and slums resulting in fundamental socioeconomic challenges (2,7,8). In 2020, slum dwellers in Africa accounted for 51.3% of urban populations, and these areas tend to be characterized by overcrowding, poor sanitation, and insufficient access to healthcare services(9,10). These factors foster environments conducive to malaria transmission, particularly in the absence of effective vector control strategies(3).

Malaria transmission is highly heterogenous across endemic countries and recent studies have reported the growing fraction of malaria burden emerging from urban areas since 2003 (2). Urban malaria control faces emerging threats such increasing insecticide resistance and the invasion of *Anopheles stephensi*, a mosquito species adept at urban breeding and increasing insecticide resistance. Moreover, urban populations tend to have lower immunity due to limited prior malaria exposure, further complicating control efforts(1). To address the stalling progress in malaria reduction in many countries, the WHO launched a global initiative in 2019 encouraging countries to stratify malaria risk based on available data and tailor interventions accordingly(11). In recognising the transmission dynamics of malaria in urban areas, WHO recently launched a global framework to respond to malaria in urban areas(12). The Ghana National Malaria Programme (NMP) has transitioned from a control program to an elimination programme in 2022(13), aiming to accelerate malaria elimination efforts by targeting interventions based on local burden. A key component of this approach was a national malaria burden stratification exercise, which identified 21 districts in the Greater Accra Region, the capital of Ghana as sustaining very low transmission and thus prioritized for malaria elimination (14).

Ghana’s stratification exercise relied heavily on malaria case reporting data from health facilities and district-level model-based malaria prevalence estimates. This approach provided important information at the district level but did not capture the fine-scale spatial heterogeneity of malaria transmission within districts. This intra-district variation in risk is relevant, particularly in urban areas where transmission is often highly focal, driven by a complex mix of socio-economic conditions, varying levels of infrastructure, and diverse landscapes(3). Understanding these micro-level variations in risk is important for guiding local malaria control efforts and ensuring that interventions are precisely targeted to areas where they are most needed. The recognition of the need for microstratification—subdividing districts into smaller, higher-risk areas for targeted interventions—has gained momentum as a strategy for improving urban malaria control in the face of limited resources and competing health demands(12). Fine-scale spatial mapping of malaria prevalence can provide the granular information necessary to identify malaria hotspots within urban areas and better understand the factors contributing to high-risk areas. This study aimed to provide a fine-scale malaria risk map for the Greater Accra Region to enhance malaria programming. Specifically, within this major urban area of Ghana, we aimed to 1) identify differences in malaria risk within districts (sub regional administrative areas, 2 determine factors associated with malaria prevalence in urban settings), and 3) provide actionable data to guide local planning for targeted intervention strategies.

## Methods

### Study area

The study was conducted in the Greater Accra Region which is home to Ghana’s national capital – Accra. It is the smallest of Ghana’s 16 administrative regions by land size but the most populous, with approximately 5,500,000 inhabitants in 2021, representing 18% of the country’s population(15). Greater Accra Region is highly urbanized with 92% of its population living in urban areas. The Region is divided into 29 administrative local government areas (LGAs), which comprise of districts, municipals, and metropolis, based on population sizes(16). The high cost of living(17) has also contributed to the development of several slum areas within the region(18).

Geographically, Greater Accra is bordered by the Eastern Region to the north, the Volta Region to the east, the Central Region to the west and the Atlantic Ocean to the south. The Region lies within the dry coastal equatorial climatic zone with temperatures between 20^0^ and 30^0^ Celsius, an ideal temperature for malaria vector and parasite development(19). Annual rainfall ranges from 635mm along the coast to 1,140mm in the northern part with two notable rainfall peaks in June and October(15). Two main rivers, the Volta and Densu, flow through the Region in addition to smaller streams that flow seasonally from the Akwapim Ridge in the Eastern Region, through lagoons, into the sea.

The Region has the lowest malaria risk out of the 16 regions in Ghana with a *Plasmodium falciparum* parasite prevalence rate of 2% according to the 2022 Ghana Demographic and Health Survey (GDHS)(20). Malaria is perennial in the region with a bimodal peak in June and October. Figure 1 shows the LGAs in the region and areas earmarked for malaria elimination by the National Malaria Elimination Program (NMEP).

**Figure 1:**
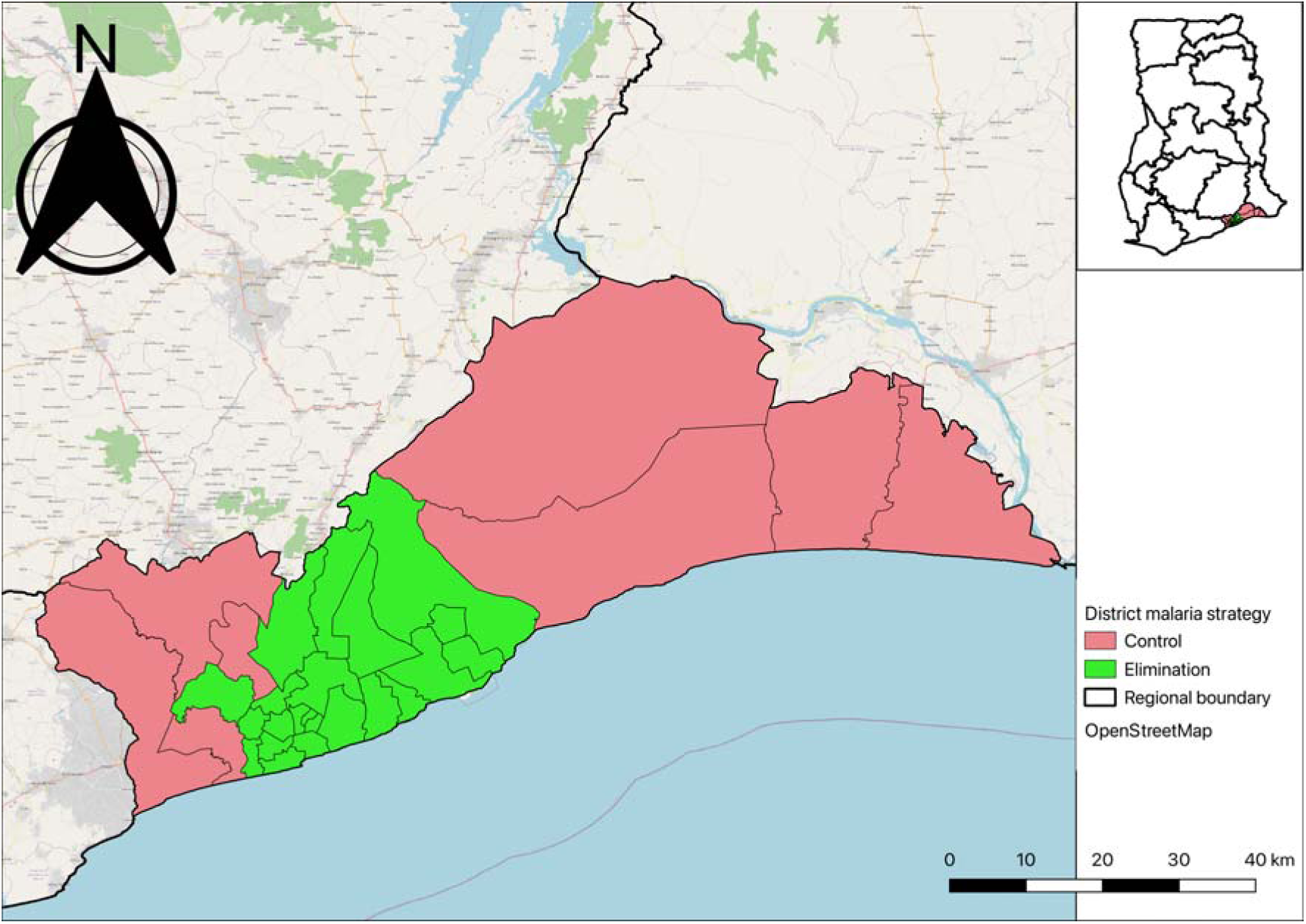
Map of Greater Accra showing districts earmarked for malaria elimination and control.

### Data

Data for this study was sourced from a cross-sectional household survey conducted across all 29 Districts of Greater Accra 2020(21) by the Kintampo Health Research Centre, Research and Development Division and the National Malaria Programme of the Ghana Health Service(22). The study population included children aged 6 months to 10 years, with the inclusion criterion of being resident in the community for at least 3 months preceding the survey. Communities and households were selected using a systematic random sampling technique. A maximum of three Subdistricts in each District were conveniently selected out of an average of four per District in the Region. This led to less populated and hard to reach areas not sampled. All communities within the selected subdistricts were listed and assigned numbers from which 14 communities were selected systematically. Within each selected community, household were selected as teams moved along directional lines, determined randomly from a central point. In each household, a maximum of two children aged 6-59 months and up to two children aged 5-10 years, eligible, were enrolled. For further details on the sampling strategy see Dosoo et al, 2024(22). The survey was conducted in October which coincides with the second peak malaria season in the region.

Trained survey teams administered a structured questionnaire to caregivers of eligible children to collect information on demographics, household characteristics and the use of malaria interventions. Additionally, each child underwent a malaria rapid diagnostic test (mRDT) specific for *Plasmodium falciparum (P. falciparum)* after receiving caregiver consent, with test results recorded as either positive or negative for *P. falciparum*. Ten percent (10%) of the sampled children were also sampled for microscopy testing for quality control purposes.

The dataset we used for this analysis included mRDT test result for each child and demographic attributes of the child such as age, sex, and place of residence. Out of the 17,035 records in the dataset, 1,378 had missing geocoordinates, while 3,257 were assigned geocoordinates at the centroid of the community they reside. Only 12,371 had coordinates at the household level and were included in the analysis (supplementary figure 1).

We obtained permission to use the data set from the National Malaria Elimination Program (NMEP) and Kintampo Health Research Centre of Ghana. The original study received ethical approvals from the Scientific Review Committee and Institutional Ethics Committee of the Kintampo Health Research Centre (KHRC).

### Patient and Public Involvement

Patient and Public were not involved in this research since it was a secondary analysis from the original study.

### Variables

The dependent variable in this analysis was *Plasmodium falciparum* infection derived from the mRDT test results, a binary outcome coded as 0 for negative and 1 for positive. Independent variables were selected based on evidence of association with malaria infection from previous studies and the availability of high-resolution raster files for the study settings(23). For this analysis, we considered 19 high-resolution remote sensing variables for this analysis and categorized them into three main broad classes – landcover, environmental/climatic, and urban (supplementary table 1). The details of how the covariates were extracted are outlined in supplementary methods section.

### Data/covariate cleaning

Data cleaning and analysis were conducted using R software, version 4.3.2(37). Covariate rasters were cropped and masked to match the shapefile boundaries of the study region. Appropriate transformations were applied to standardize the rasters. The mean values of the rasters were then extracted for all points in the dataset for analysis.

### Exploratory analysis

We conducted an exploratory analysis to understand the relationships between the malaria prevalence and the suit of explanatory covariates. We first performed a univariate logistic regression analysis to identify covariates significantly associated with the dependent variable. Additionally, a non-spatial multivariate logistic regression model was used to assess the combined effect of all covariates on the dependent variable. We excluded crops and flooded vegetation as they did not show any informative association with the dependent variable. We also plotted point coordinates on the map of the study area to visualize the spatial distribution of the RDT test results.

### Variable selection

The inclusion of urban covariates at 100m resolution was to allow us to consider the landscape heterogeneity we find in typical urban areas in Africa in our prediction. In order not to lose the full effect of the urban covariates in our model, we combined their effect through a principal component analysis (PCA). The PCA enabled us to transform the high-dimensional, correlated urban covariates into a low-dimensional space enabling us to capture the relevant information we need(38). We used the *rasterPCA()* function in R(39) for the PCA analysis, selecting four principal components (PC1, PC2, PC3, and PC4) that explained approximately 99% of the variance in the data. PC1-4 replaced the five urban covariates in the suite of 19 covariates considered for variable selection.

We then tested for multicollinearity among the covariates using the variable inflation function (VIF) and sequentially dropped covariates with a VIF over 10 until all remaining covariates had a VIF <10. This led to the removal of NDVI and LST band 10, and TCG with 14 covariates retained. See supplementary table 2 for the final set of covariates that were included in the model fitting.

### Geostatistical model

The geostatistical model used in this analysis assumed that the probability *p* of a child *x_i_* testing positive for *Plasmodium falciparum* malaria at a location *i =1, …, n* follows a binomial distribution, expressed as:

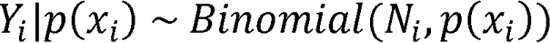

Where *Y_i_* represents the number of children testing positive at a location *i* out of *N_i_* children tested. The outcome of every test was binary, positive, or negative for *Plasmodium falciparum* malaria.

The binomial geostatistical Model is formulated using the logit transform as:

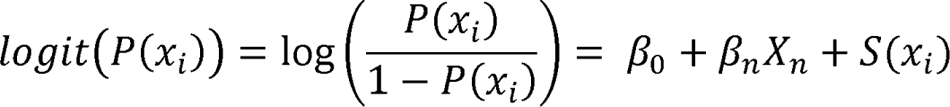

where β_0_ is the intercept parameter, β_n_ is the vector of covariate coefficients for the covariates in the design matrix *X_n_*, and *S(x_i_)* is the spatial random field, which represents the spatial variation between the sampled locations. We assumed the spatial random field is a zero-mean Gaussian process with Matérn covariance function. The covariance between the two points located at *x_i_* and *x_j_* is given by;

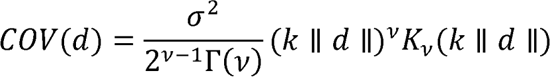

where *d* is the distance between location *x_i_* and *x_j_*; σ^2^ is the variance; ν is the smoothness parameter that determines the analytic smoothness of the process; *K_V_*(.) is the modified Bessel function of second kind and order *v* > 0; *k* > 0 is the range parameter which determines how quickly the covariance decays as distance increases; and Γ(*v*) is the Gamma function.

### Model fitting

The model for this analysis was fit using the Integrated Nested Laplace Approximations (INLA) package in R, version 24.02.09. The R-INLA package has been shown to be a computationally efficient alternative(40,41) to the Markov chain Monte Carlo (MCMC) method, particularly when combined with the Stochastic Partial Differential Equation (SPDE) approach(42). The combination of INLA and SPDE facilitates efficient point-level data analysis within a Bayesian framework(43,44). To begin, we constructed a mesh surface across the boundary of the study region and built an SPDE model to generate the index set. We also built a projection matrix that will project the mesh, on which the spatial random field is represented, to the observed locations. Finally, we combined the data, effects, and projection matrix into a stack object for the model fit.

We used the default priors in R-INLA for the hyperparameters in this analysis, namely: the variance and range. We fitted the model in R-INLA using the selected covariates and a spatial random field to predict the probabilities of a child testing positive at each observed location. We explored statistical significance to malaria parasite infection by extracting the exponentiated covariates slopes.

### Model validation

The sampling method for the data resulted in some parts of the region not sampled and thus, without data leading to possible bias in prediction estimates. We therefore, employed two approaches to evaluate the model: 1) we randomly partitioned the data into 10-folds, with each fold having similar number of observations, 2) we used spatial block cross-validation (CV)(45) to split the data randomly into 10-folds over the study area with each fold having different number of observations for the test and training data (supplementary table 3). These two approaches were to enable us to assess 1) how well our model fitted the data overall and 2) how well our model predicted the data in both sampled and unsampled areas. In the case of the random cross-validation, the model was trained 10 times. In each iteration, one-fold was held as a test set and the other nine were used as training set. In the spatial cross-validation, a set of data in particular blocks within each fold were randomly allocated as test and training sets for model prediction and model fitting respectively. In both approaches, we blinded the response variable in the test data by setting its value to ‘NA’ so that it did not use observed data for prediction. This step was repeated ten times, changing the fold ID in each iteration. We assessed the model performance by calculating the root mean squared error (RMSE) for each iteration for the model prediction and the model fitting (which we tagged as estimation). We then compared the computed RMSEs for the two approaches to make a judgement on the model performance. RMSE is an indication of the average deviation of the predictions from the actual observations and is one of the recommended metric for evaluating prevalence models(46). A lower RSME is an indication of a model’s ability to predicted the observed data well(47).

### Model prediction

To explore spatial variation in malaria prevalence across the region and predict in unsampled areas, we first extracted spatial points from one of the covariate raster to get a spatial grid surface of 100m resolution. This resulted in 434,280 points for prediction. We then extracted 100 samples from the posterior of the full fitted model to predict malaria prevalence for children 6 months to 10 years at 100m for the study area. We calculated mean predicted probabilities, standard deviations (SD), and credible intervals (CI) for each pixel and generated maps for the mean prediction and its associated SD, and CI.

### Exceedance probability

To assess the probability that the prevalence of malaria in some areas within the region exceeded 10%, we conducted an exceedance probability (EP) analysis using a 10% threshold. The 10% threshold was chosen to align with the WHO classification of <10% parasite prevalence for low transmission settings (48,49). The EP account for the level of uncertainty around our prediction and provides useful information for making decisions. In this study, the EP was used to identify areas within low transmission areas that are likely to have malaria prevalence above 10% and therefore not appropriate to be targeted for malaria elimination interventions by the NMEP. To calculate the EP, we assumed the cumulative distribution function (CDF) of the predictions was a beta distribution with values constrained between 0 and 1. Thus the EP was calculated as

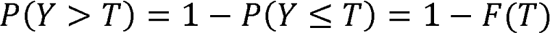

where *P(Y > T)* is the exceedance probability of predicted prevalence *Y* exceeding a threshold *T* and *F(T)* is the CDF of the predicted values. We derived the alpha and beta shape parameters from the posterior means and standard deviation of the predictions. The alpha and beta parameters controls the shape of the distribution and sets them between 0 and 1 respectively(50).

## Results

### Descriptive analysis results

The final data of 12,371 children aged 6 months to 10 years had more females (53%) than males (47%). The number of children who tested positive for mRDT were 536 (0.04%) while 11,835 (95.6%) tested negative. More of the children (51%) were aged 6month to 5 years. mRDT positivity was significantly higher in children 5-10 years (59%) compared with children less than 5 years (41%). Out of the 6,333 children for which place of residence were recorded, majority (68%) lived in an urban area and the mRDT results between the different areas of residence was statistically significant (p-value <0.001). Proportionally, children residing in urban areas had higher mRDT positive results (46%) compared with children in peri-urban (24%) and rural areas (30%). Table 1 highlights the demographic factors and RDT results in the dataset.

**Table 1:** Demographic parameters of study participants and their RDT results.

| Variable | N | RDT results |  |  |  |
| --- | --- | --- | --- | --- | --- |
|  |  | Overall, N =<br>12,371<sup>1</sup> | Negative, N =<br>11,835<sup>1</sup> | Positive, N =<br>536<sup>1</sup> | p-value<sup>2</sup> |

| RDT results |  |  |  |  |  |
| --- | --- | --- | --- | --- | --- |
| Variable | N | Overall, N =<br>12,371<sup>1</sup> | Negative, N =<br>11,835<sup>1</sup> | Positive, N =<br>536<sup>1</sup> | p-value<sup>2</sup> |
| Sex | 12,371 |  |  |  | 0.3 |
| Female |  | 6,548 (53%) | 6,276 (53%) | 272 (51%) |  |
| Male |  | 5,823 (47%) | 5,559 (47%) | 264 (49%) |  |
| Age group | 12,371 |  |  |  | <0.001 |
| 0<5 |  | 6,370 (51%) | 6,150 (52%) | 220 (41%) |  |
| 5-10 |  | 6,001 (49%) | 5,685 (48%) | 316 (59%) |  |
| Place of residence | 6,333 |  |  |  | <0.001 |
| Rural |  | 968 (15%) | 889 (15%) | 79 (30%) |  |
| Peri_urban |  | 1,068 (17%) | 1,004 (17%) | 64 (24%) |  |
| Urban |  | 4,297 (68%) | 4,175 (69%) | 122 (46%) |  |
| Unknown |  | 6,038 | 5,767 | 271 |  |

<sup>1</sup>n (%)
<sup>2</sup>Pearson's Chi-squared test

### Exploratory analysis results

#### Univariate and multivariate analysis

Supplementary table 4 highlights the output of the univariate and multivariate analysis conducted to determine the association between the outcome variable (mRDT positive) and the explanatory variables. In the univariate regression analysis, age group and place of residence i.e., living in an urban area, were found to be significantly associated with malaria prevalence. Among the land cover covariates, water and tree coverage were significantly associated with malaria prevalence. Land surface temperature bands 10 (LST_10) and 11 (LST_11), enhanced vegetation index (EVI), normalized difference vegetation index (NDVI), tasselled cup greenness (TCG), and tasselled cup wetness (TCW) were the climatic covariates significantly associated with the odds of malaria prevalence. All the urban covariates - settlement characteristics (built_c), building height (built_h), built surface (built_s), building volume (built_v), and population density (pop), were found to be significantly associated with the outcome. In the multivariate regression analysis, age group (5-10 years), place of residence (urban), LST_10, TCB, TCG and Pop significantly influence the odds of malaria prevalence in the region.

We mapped the the distribution of mRDT results over the study area (supplementary figure 2). We observed that data was more clustered in the districts earmarked for elimination due to their population size compared with districts located outside the elimination.

### Principal Component Analysis (PCA) results

The result of the PCA we conducted on the five urban covariates are shown in supplementary table 5. While PCA1 explained 86.6% of the variation in the covariates, we included PCA1-4, which cumulatively accounted for almost 100% of the variation.

### Geostatistical model results

The geostatistical model results showed that while predictors such as TCB and PC3 were significantly associated with reduced odds of malaria prevalence, EVI and PC2 were associated with increased odds (Table 2). TCB was associated with a reduction in odds of malaria prevalence by 36% (OR-0.64, CI – 0.47,0.85) while PC3 was associated with a reduction of 18% (OR – 0.82, CI – 0.72,0.93). Conversely, EVI and PC2 were associated with an increase in odds of malaria prevalence by 47% (OR – 1.47, CI – 1.14,1.88) and 13% (OR – 1.13, CI – 1.03,1.25) respectively. Covariates such as water, trees, bare ground, rangeland, LST_11, TCW, PC1, and PC4 were found not to significantly affect the odds of malaria prevalence in children in the Greater Accra region.

**Table 2:** Odds ratios of posterior mean of model parameters including standard deviation, lower and upper credible intervals.

| Parameter | Mean | Standard deviation | Lower CI | Upper CI |
| --- | --- | --- | --- | --- |
| intercept | 0.07 | 1.35 | 0.04 | 0.13 |
| Water | 1.06 | 1.22 | 0.72 | 1.55 |
| Trees | 1.09 | 1.16 | 0.81 | 1.46 |
| Bareground | 1.14 | 1.18 | 0.83 | 1.57 |
| Rangeland | 0.96 | 1.14 | 0.74 | 1.25 |
| LST_11 | 0.83 | 1.12 | 0.66 | 1.05 |
| EVI | 1.47 | 1.13 | 1.14 | 1.88 |
| TCB | 0.64 | 1.16 | 0.47 | 0.85 |
| TCW | 0.71 | 1.20 | 0.50 | 1.01 |
| PC1 | 1.02 | 1.04 | 0.95 | 1.10 |
| PC2 | 1.13 | 1.05 | 1.03 | 1.25 |
| PC3 | 0.82 | 1.07 | 0.72 | 0.93 |
| PC4 | 1.02 | 1.16 | 0.75 | 1.37 |
| Range | 0.08 | 1.60 | 0.03 | 0.20 |
| Variance | 0.51 | 1.41 | 0.27 | 1.02 |

### Model validation

The average RSME for the final model was 0.199 (measured in the same units as the dependent variable – i.e., malaria infection prevalence expressed as a fraction between zero and 1). Supplementary figure 3 shows results of the spatial blocking of the study area into folds where data in some blocks we assigned as either test or train set.

We compared the RMSE of the prediction and estimation models for both random and spatial blocking cross validations for each fold (supplementary table 6). Generally, we expect RMSE values from the two models to be either similar or very close. From supplementary table 6, we observed that the prediction RMSEs are quite close to the estimation RMSE in the random cross validation. In some of the folds (2, 5, 6, 8, 9 and 10) we observed lower RSME for the prediction model than the Estimation model. In the spatial blocking cross validation, we observed a similar trend. In four folds (2, 4, 5 and 6), however, we noted much higher differences in the prediction model than the estimation model. Overall, the RSMEs from the two approaches were closer to average RSME of the final model.

### Prediction

We produced a prediction map of the malaria parasite prevalence in children under 10 years in Greater Accra region at 100m resolution. The predicted malaria prevalence ranged from 0.003 to 0.517 (supplementary figure 4). We rescaled and re-categorized the estimated malaria prevalence into operationally meaningful cut-offs (less than 1%, 1-5%, 5-10%, 10-25%, >=25%) to align with thresholds used to stratify malaria risk in the National Malaria Elimination Strategic Plan. We then overlaid the prevalence map with district administrative boundaries to generate malaria prevalence map at district level (Figure 2). From the map, we observe lower malaria prevalence in the central part of the region compared with areas in the northern and eastern parts of the region. The northern part of the region which borders the Eastern region of Ghana and the south-eastern part of the region, have predicted parasite prevalence above 10% with some areas showing prevalence above 25%. Districts within the pre-elimination earmarked areas were found to have malaria parasite prevalence below 5% while districts outside that zone showed prevalence mostly above 5% as highlighted in the insert map. Whiles most of the pre-elimination areas showed prevalence below 5%, the northern part of districts like Ayawaso West, Ga East, La-Nkwantanang Madina and Adenta had prevalence above 10%.

**Figure 2:**
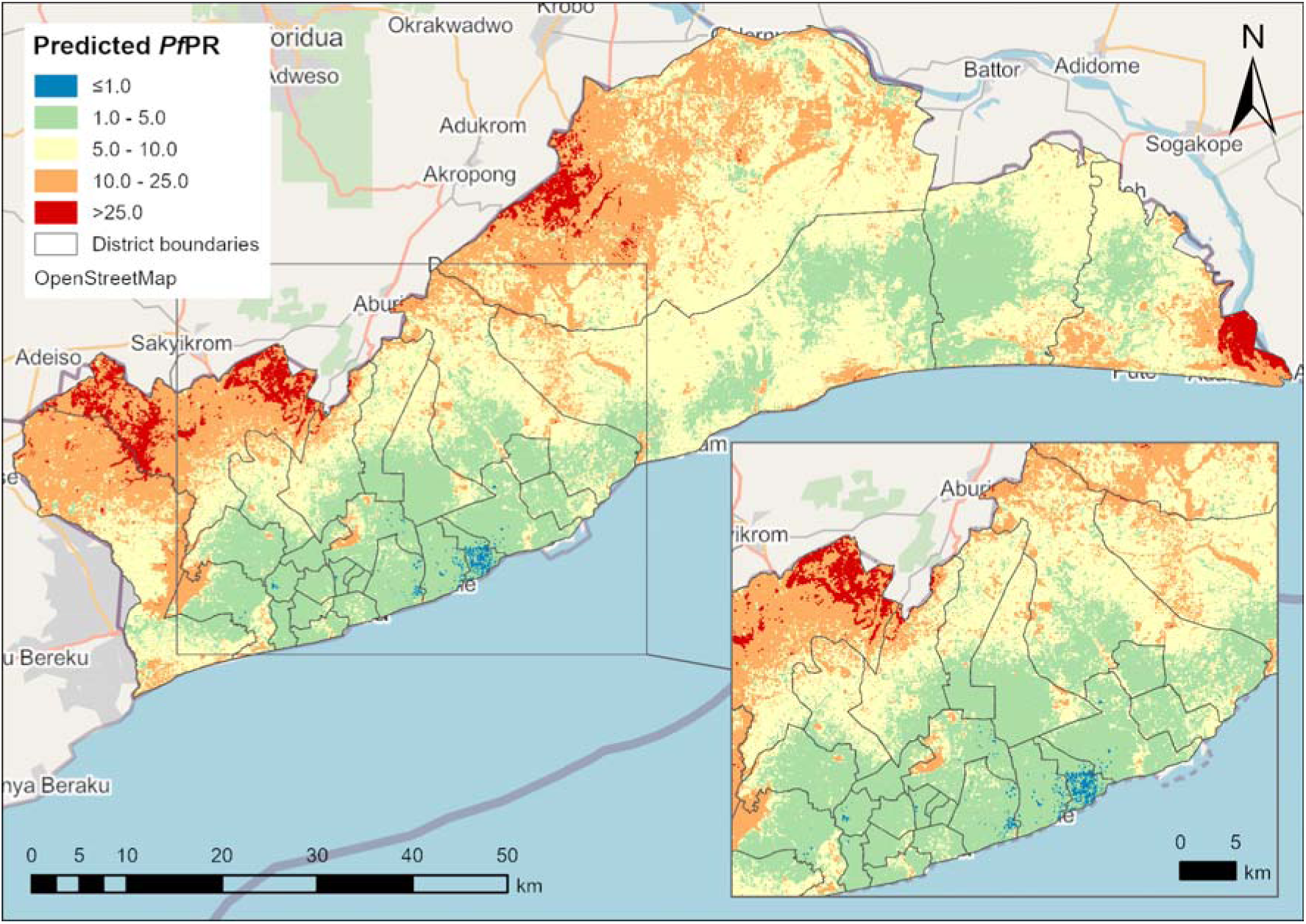
Predicted mean malaria parasite prevalence in Children 6month-10 years in Greater Accra region. Insert, predicted mean malaria prevalence in Districts earmarked for elimination.

Supplementary figure 5 is a map of the prediction standard deviation, which quantifies the degree of uncertainty around our estimate of the parasite prevalence for the region. The standard deviation map showed that the variation in our prediction was between 0 and 24%. Overall, the uncertainty of our prediction is low.

### Mapping exceedance probabilities

We calculated the exceedance probabilities of malaria prevalence being greater than 10% (Figure 3). The areas close to 0 on the scale, are more likely to have malaria prevalence below 10% and should be targeted for elimination activities. The areas close to 1 are thus, more likely to have prevalence above 10% and should be targeted for control interventions.

**Figure 3:**
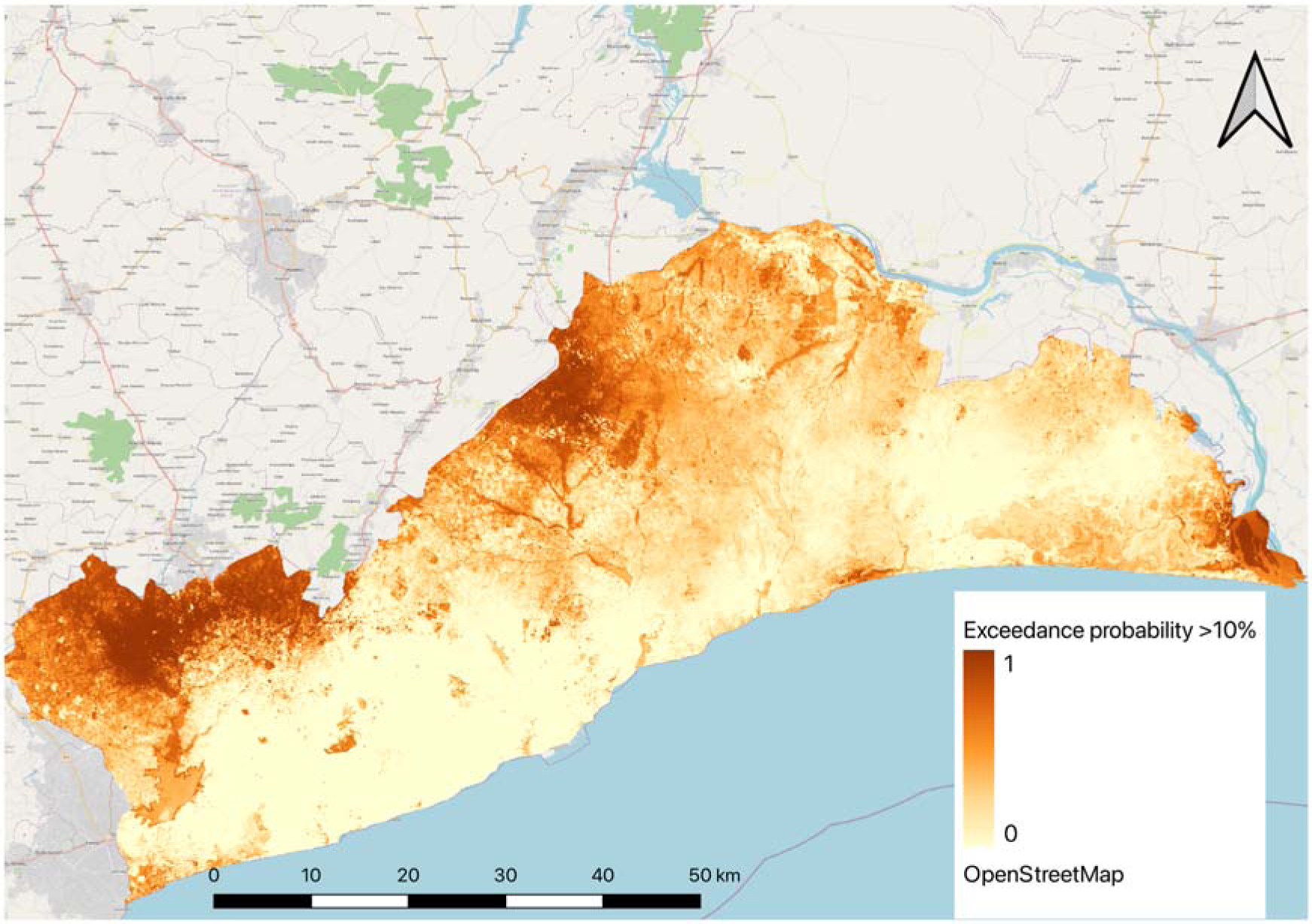
Exceedance probability of malaria parasite prevalence above 10% in Children 6month-10years in Greater Accra region

## Discussion

The study sought to achieve three specific objectives; 1) identify differences in malaria risk within and among districts, 2) explore factors associated with malaria prevalence in the region, and 3) provide information to better guide local planning of malaria interventions to deploy within the same district. The data collection coincided with a major raining season, allowing us to assess a period of peak malaria risk in the region. Moreover, every District in the region was sampled, making the predictions comparable and representative.

For our first objective, we predicted malaria prevalence for children 6 months to 10 years and younger to be as low as 0.3% and as high as 52%. The West-central part of the region right through to the coastal and middle part of the region showed relatively low malaria prevalence compared with the upper/ forest areas. The coastal and south-central part of the region are also more urbanized with high population density. This finding tends to agree with other studies which have reported lower malaria risk in urban areas than peri-urban and rural areas(3,51). As found by Kabaria et. al., 2016, malaria risk within urban areas is heterogenous(52). The heterogeneity of malaria within the highly urban areas was evident in our prediction map where we observed malaria prevalence ranging from <1 to 25% in the areas earmarked for pre-elimination. The high prevalent areas within the highly urbanized part of the region have been identified as slum areas according to MacTavish et.al(18). This may be attributed to some species of mosquito like the *An. gambiae* are which are increasingly adapting to polluted breeding sites in urban areas(2).

We observed that, predictions at 100m spatial resolution as used in our study, enabled us to capture finest spatial variations in neighbourhoods compared with similar studies that predicted at 1km or 5km(23,51–53). In rapidly growing urban settings in sub-Saharan Africa where there are profound differences in topography and socioeconomic factors, use of household or individual level data becomes imperative to capture difference in risk. The use of individual level instead of cluster level data, enabled us to make accurate estimates that accounts for these differences. Inferences at this lowest level become prohibitive when using cluster level data from DHS/MIS surveys where cluster locations are displaced by up to 5km in rural clusters and up to 2km in urban clusters(54). This level of displacement can infer significant bias(55) especially in urban settings where malaria is highly focal and there can be incredible variability in malaria risk in short distances(3,52).

While prevalence studies predict malaria risk in asymptomatic population, malaria incidence reported from health facilities gives an indication of malaria illness in the infected population. Two studies used the latter to predict malaria risk in the same study region(51,56). They observed seasonal pattern in malaria cases where it peaks in June/July, drops in September, and increases in October. The districts we identified as having high malaria prevalence above 10% aligns with the findings from these studies. Our work therefore compliments previous studies in the region and provides vital insights on malaria risk in the region using incidence and prevalence.

Model validation results show that we were able to predict malaria prevalence in Greater Accra with a high level of accuracy, evident from the low RMSE values we obtained in all the iterations in the two approaches. Additionally, the closeness of the prediction RMSE to the estimate RMSE, even though our test set did not include the observed values, indicates the model was able to predict well in unsampled areas. The wide age group from our data, 6 months to 10 years, enabled us to profile malaria risk in the most at risk group as there have been reported shift in severe malaria in most sub-Saharan countries from children under five to older children as a result of chemoprevention strategies targeted at children aged less than 5 years(62).

Assessing factors associated with malaria prevalence in Greater Accra, we found four covariates to be significantly associated with the odds of a child testing positive for malaria. Enhance vegetation index was the environmental covariate found to increase the odds of malaria prevalence. This suggests that areas with a lot of vegetation tend to create conducive environment for breeding of mosquitoes, consistent with other studies in Kenya(57) and Burkina Faso(58). High tasselled cap brightness values –usually associated with partially covered soil or features such as concrete or asphalt, was found to reduce the odds of malaria prevalence. Covered, concrete or asphalted surfaces tend to provide very limited opportunity for breeding of mosquitoes. PC3 was found to reduce the odds of malaria prevalence by 18% whiles PC2 increases the odds by 13%. Though we couldn’t identify the individual urban covariates that influences malaria prevalence, results of the non-spatial multivariate regression suggests that areas with high population density have reduced the odds of malaria prevalence. This finding is supported by Kabaria et al (2017), who found that malaria risk curve significantly reduces in areas with population densities >1000 persons per km^2^(59). Factors explaining PC2 association with increased odds of malaria prevalence could not be explained in this study and will therefore need further research.

This study had a number of important limitations. The study did not account for temporal or seasonal variations in malaria risk in the region. Even though data collection for the original study coincided with the second peak season of malaria in the region, it will have been ideal if it captured the entire malaria transmission season to enable programme managers to consider optimal intervention timing. The covariates used in this analysis were extracted for 2020, the year of the survey. Accra, like many urban areas in Africa is rapidly expanding with high population migration due to economic activities and therefore frequent changes in topography occur. Health system intervention like access to health services has improved since 2020 with expansion of health insurance as well as improvements and/or increase in health infrastructure. Increase in the number of health facilities and improvement in primary health services leading to reduced travel time and good access to care have been found to reduce malaria disease burden(60,61). There will be the need to repeat this kind of analysis using more recent survey data in scope and form and remote sensing data that reflect the current landscape in urban areas in the region. Additionally, the analysis did not include household level factors such as type of building material used for the walls, floors, and ceiling as well as type of malaria intervention used in the household. We are of the view that the addition of household factors will help improve our understanding of the factors influencing the heterogeneity we see within highly urbanised areas as found in studies conducted elsewhere(3).

The study results have provided useful information to support decision making at both national and subnational levels on deployment of strategic malaria interventions. The observed range of prevalence Greater Accra region has implications for malaria interventions that should be deployed. While the latest DHS survey estimated a prevalence of 2% for the region(20), our study has identified districts and areas with prevalence as high as 52%. From our results studies we believe there is an epidemiological shift in the region, in line with the findings of Dosoo et al,2024(22) and therefore interventions targeting older children should be deployed in the region. The exceedance probability map developed show areas within each district that are likely to have malaria prevalence above 10% and therefore needs control interventions. With the call by WHO for countries to stratify malaria risk and tailor interventions to where they are need most, our analysis provides a blueprint for the NMEP and district health managers to implement strategies to reduce malaria prevalence in high-risk areas.

## Conclusion

Our study has shown that, crops, tasselled cap wetness and urban covariates like building volume and population density reduces the odds of malaria prevalence among children 6 months to 10 years in Greater Accra region. This work has identified the spatial heterogeneity of malaria in the region. The northern part of the region and south-eastern most part of the region has higher malaria risk compared with the central part. Within the districts earmarked for pre-elimination, we identified hotspots where the probability of malaria prevalence above 10% is greater and therefore remains a threat to the elimination agenda. These areas need to be targeted for control interventions to drastically reduce the malaria burden.

Additionally, this work will hopefully be a guide for the NMEP and health service managers in the district and the region to target the right intervention to a particular area based on the level of malaria risk. This will enhance better use of scarce resources to achieve maximum impact.

## Supplementary information

### Figures

Supplementary figure 1: Cascade of final number of observations used in the analysis.

Supplementary figure 2: Distribution of RDT results of study participants

Supplementary figure 3: Spatial assignment of data into 10-folds for cross validation

Supplementary figure 4: Smooth predictive map of malaria prevalence for children 6 month to 10 years in Greater Accra Region

Supplementary figure 4: Standard deviation of predicted malaria prevalence in Children 6month-10years in Greater Accra region

#### Tables

Supplementary table 3: Description of covariates, source, and spatial resolution

Supplementary table 2: Final set of covariates and their VIF values

Supplementary table 3: Number of observations for each fold for random and spatial blocking cross-validations

Supplementary table 4: Univariate and Multivariate regression results

Supplementary table 5: PCA results showing the relative PCA components.

Supplementary table 6: Root Mean Squared error (RMSE) of prediction and estimation models for random and spatial blocking cross validations.

#### Ethics approval

The original study was approved by the Ethical review boards of the Ghana health service. Anonymized data for this work was obtained from the Kintampo Health Research Centre through the Ghana National Malaria Elimination Programme. This study is within a PhD projected under the Malaria Atlas Project with ethical approval from the Ethics committee of Curtin University – HRE2021-0734.

## Data Availability

The data that support the findings of this study are available on reasonable request, subject to ethics, governance, and privacy considerations. The code for the analysis is available on the personal GitHub portal of the corresponding author and will be made available when necessary.

## Acknowledgements

We acknowledge the contribution of staff of the Kintampo Health Research Centre who undertook the main study and collected the data for this analysis. we also acknowledge the efforts of staff of the National Malaria Elimination Programme, Regional and District Health Directorates of the Ghana Health Service in Greater Accra Region, as well the respondents of the main study.

## Authors contribution

S.K.O, P.A, K.A.A., and P.G. conceptualized this study.

DKD, NYP, KPA and KLM conceptualized and developed the protocol for the original study.

DKD, GAA, WM, KK, and KPP coordinated the study, trained, and supervised field teams and quality assured data for analysis.

SKO, PA, KAA, PG and KLM refined the analysis plan for this study.

SKO and JR extracted and processed the spatial covariates used in the analysis.

PA, MM and SKO reviewed and refined the model used for the analysis.

SKO conducted the analysis, developed the model, and wrote the first draft of the manuscript.

All authors contributed to the interpretation of the results of this study. All authors critically reviewed this manuscript, made inputs, and approved the final submitted version.

## Materials & Correspondence

All correspondence and material requests should be addressed to Samuel Kweku Oppong

## Copyright

The Corresponding Author has the right to grant on behalf of all authors and does grant on behalf of all authors, an exclusive licence to the BMJ Publishing Group Ltd to permit this article (if accepted) to be published in BMJ editions and any other BMJPGL products and sublicences such use and exploit all subsidiary rights, as set out in our licence.

## Transparency declaration

The lead author affirms that the manuscript is an honest, accurate, and transparent account of the study being reported; that no important aspects of the study have been omitted; and that any discrepancies from the study as planned have been explained.

