## Supplementary methods for "Fine scale spatial mapping of urban malaria prevalence for microstratification in an urban area of Ghana"

Supplementary method section

Covariate extraction

We derived global images for all variables, except the urban variables, from Landsat 8 data at 30m spatial resolution and aggregated them to 100m resolution using Google Earth Engine(24). To create cloud-free images, we employed the Earth Engine Simple Composite algorithm, which selects the 10 least cloudy pixels for each location, generating a median value. This process was performed for each of the four seasons, and a yearly average was then computed to minimize the influence of seasonally biased cloud cover.

Seven land cover classes were selected based on their relevance to the study area's topography. These classes include water, trees, crops, built area, rangeland, bare ground, and flooded vegetation. For each class, we calculated the proportion of the pixel covered by that specific land cover over the study area boundaries.

For environmental and climatic variables, we included Tasselled Cap Brightness (TCB), Tasselled Cap Wetness (TCW), Tasselled Cap Greenness (TCG), Enhanced Vegetation Index (EVI), Normalised Difference Vegetation Index (NDVI) and Land surface temperature (LST). EVI and NDVI were calculated using standard formulas:

- $EVI=2.5*\left( \frac{Band5-Band4}{Band5+6\times Band4-7.5 \times Band2+1} \right)$(25)
- $NDVI =\frac{\left( Band 5 - Band 4 \right)}{\left( Band 5 + Band 4 \right)}$ (26)

TCB, TCW, TCG were derived using coefficients from Baig, M. H. A., et al. (2014) (27). LST, which influences the rate and timing of plant growth (28) was calculated using the simplified version of the Planck's law-based LST calculation that is commonly used in remote sensing and thermal imagery analysis (29).

$LST=\frac{B_{T}}{(1+w\left( \frac{B_{T}}{p} \right)*\ln\left( e \right))}$

where$B_{T}$ is Brightness Temperature (measured from thermal infrared remote sensing data)$w$ is wavelength of the emitted radiance,$p$ is Planck's constant, and $e$ is Emissivity (surface emissivity). We calculated LST separately for bands 10 (LSTemp_10) and bands 11 (LSTemp_11) from Landsat and included emissivity from MODIS11A2 v061 using band 31 and band 2 for LSTemp_10 and LSTemp_11 calculations respectively (30).

We obtained the urban variables from the Global Human Settlement Layer (GHSL) website(31). The GHSL data included GHS population grid(32), GHS built-up volume grid(33), GHS built-up surface grid(34), GHS settlement Characteristics(35), and GHS building height grid(36).
