## Supplementary figure 5 for "Fine scale spatial mapping of urban malaria prevalence for microstratification in an urban area of Ghana"

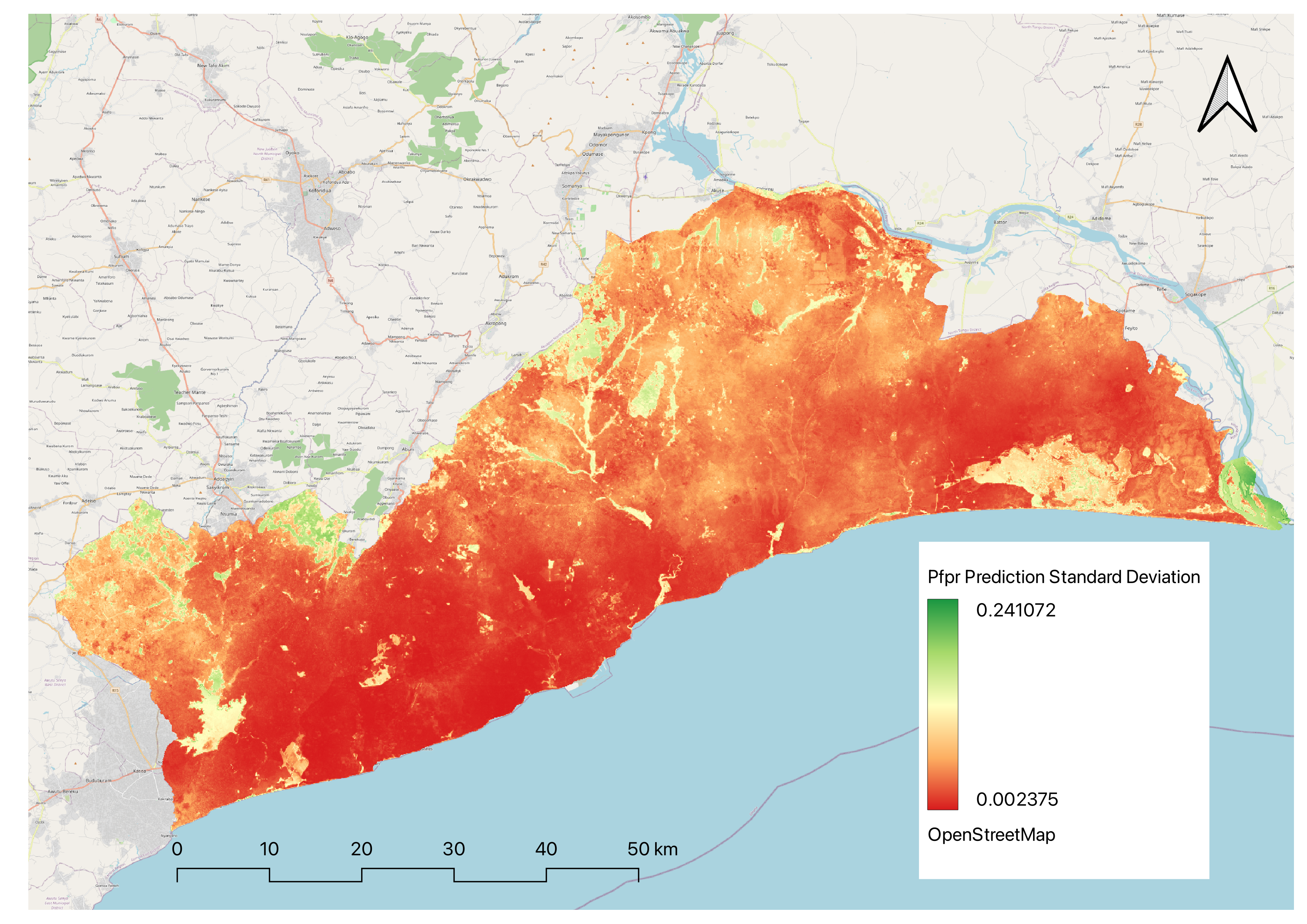

Supplementary figure 5: Standard deviation of predicted malaria prevalence in Children 6month-10years in Greater Accra region
