## Supplementary figure 4 for "Fine scale spatial mapping of urban malaria prevalence for microstratification in an urban area of Ghana"

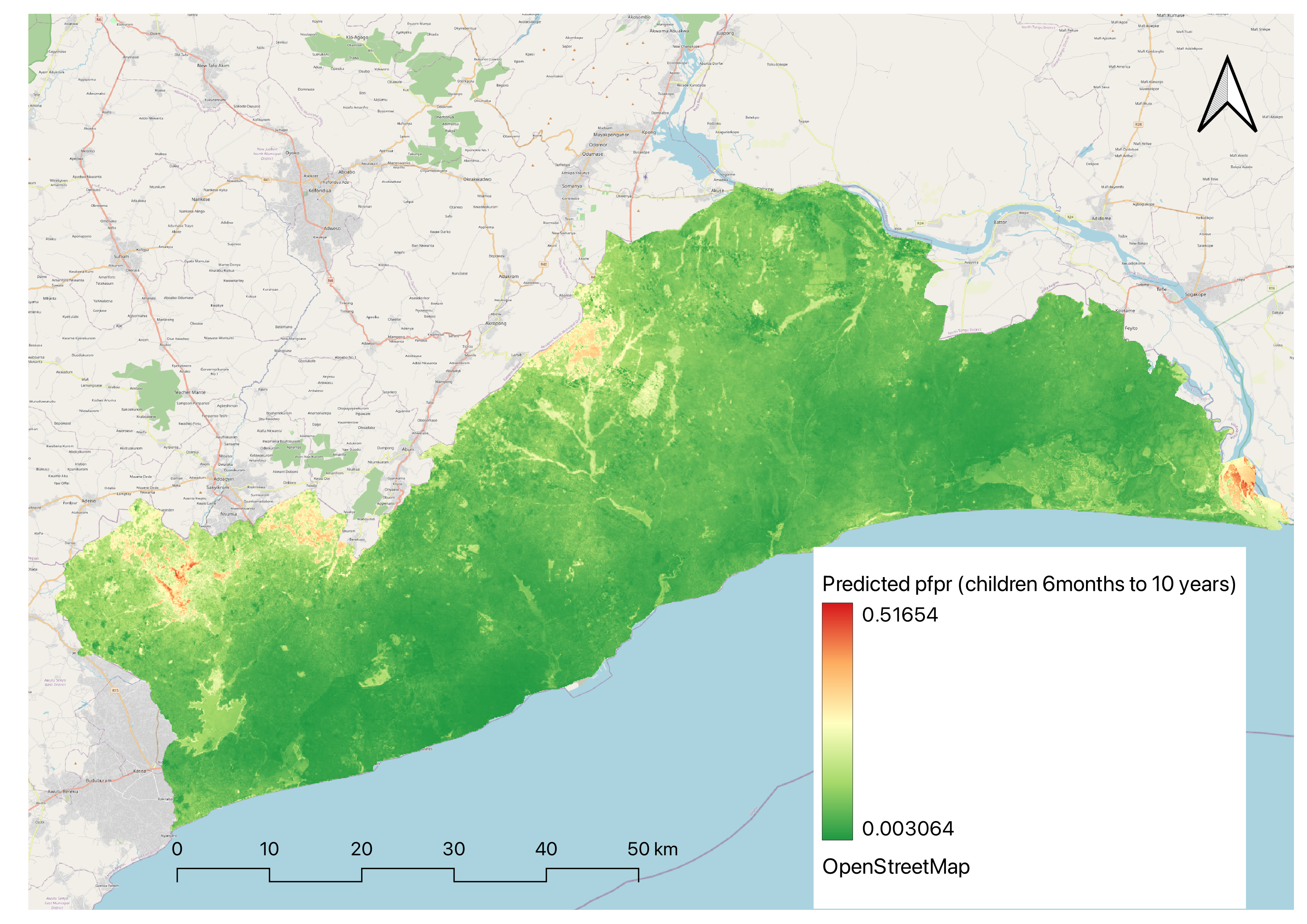

Supplementary figure 4: Smooth predictive map of malaria prevalence for children 6 month to 10 years in Greater Accra Region
