## Supplementary table 6 for "Fine scale spatial mapping of urban malaria prevalence for microstratification in an urban area of Ghana"

Supplementary table 6: Root Mean Squared error (RMSE) of prediction and estimation models for random and spatial blocking cross validations.

| Random cross validation | Spatial blocking cross validation |
| --- | --- |
| Fold \| Prediction RMSE \| Estimation RMSE; 1 \| 0.2180 \| 0.1970; 2 \| 0.1863 \| 0.2006; 3 \| 0.2397 \| 0.1942; 4 \| 0.2075 \| 0.1984; 5 \| 0.1940 \| 0.1999; 6 \| 0.1763 \| 0.2017; 7 \| 0.2085 \| 0.1982; 8 \| 0.1828 \| 0.2010; 9 \| 0.1982 \| 0.1993; 10 \| 0.1871 \| 0.2006 | Fold \| Prediction RMSE \| Estimation RMSE; 1 \| 0.1941 \| 0.2007; 2 \| 0.3020 \| 0.1955; 3 \| 0.1821 \| 0.2031; 4 \| 0.2980 \| 0.1974; 5 \| 0.3466 \| 0.1937; 6 \| 0.3024 \| 0.1946; 7 \| 0.2410 \| 0.1952; 8 \| 0.1540 \| 0.2177; 9 \| 0.2185 \| 0.1984; 10 \| 0.1788 \| 0.2002 |
