## Supplementary table 5 for "Fine scale spatial mapping of urban malaria prevalence for microstratification in an urban area of Ghana"

Supplementary table 5: PCA results showing the relative PCA components. Table shows the standard deviation, the proportion of variation and cumulative proportion for each principal component.

| PCA components | PCA1 | PCA2 | PCA3 | PCA4 | PCA5 |
| --- | --- | --- | --- | --- | --- |
| Standard deviation | 2.042 | 0.597 | 0.499 | 0.201 | 0.024 |
| Proportion of variation | 0.866 | 0.074 | 0.052 | 0.008 | 0.000 |
| Cumulative Proportion | 0.866 | 0.9340 | 0.991 | 0.999 | 1 |
