## Supplementary figure 1 for "Fine scale spatial mapping of urban malaria prevalence for microstratification in an urban area of Ghana"

Supplementary figure 1: Cascade of final number of observations used in the analysis.

17,035 children 6months to 10 years enrolled.

29 records without RDT results

17,006 records with RDT results

15,628 records with geocoordinates

1,378 records without geocoordinates

12,371 records with household level geocoordinates

3,257 records with geocoordinates for the centroid of the community
