## Supplementary table 4 for "Fine scale spatial mapping of urban malaria prevalence for microstratification in an urban area of Ghana"

Supplementary table 4: Univariate and Multivariate regression results. Table shows the number of observations for each variable, the odds ratio (OR), lower and upper credible intervals (95% CI) and the p-value for both univariate and multivariable regression.

|  | **Univariate** | | | | **Multivariable** | | |
| --- | --- | --- | --- | --- | --- | --- | --- |
| **Characteristic** | **N** | **OR**^1^ | **95% CI**^1^ | **p-value** | **OR**^1^ | **95% CI**^1^ | **p-value** |
| Sex | 12,371 |  |  |  |  |  |  |
| Female |  | — | — |  | — | — |  |
| Male |  | 1.10 | 0.92, 1.30 | 0.3 | 1.06 | 0.82, 1.36 | 0.7 |
| Age group | 12,371 |  |  |  |  |  |  |
| 0<5 |  | — | — |  | — | — |  |
| 5-10 |  | 1.55 | 1.30, 1.85 | <0.001 | 1.45 | 1.12, 1.87 | 0.004 |
| Place of residence | 6,333 |  |  |  |  |  |  |
| Rural |  | — | — |  | — | — |  |
| Peri_urban |  | 0.72 | 0.51, 1.01 | 0.057 | 0.76 | 0.50, 1.15 | 0.2 |
| Urban |  | 0.33 | 0.25, 0.44 | <0.001 | 0.59 | 0.40, 0.89 | 0.011 |
| Water | 12,341 | 1.71 | 1.20, 2.33 | <0.001 | 1.11 | 0.68, 1.72 | 0.6 |
| Trees | 12,341 | 1.95 | 1.50, 2.50 | <0.001 | 1.10 | 0.75, 1.61 | 0.6 |
| Built_area | 12,341 | 0.91 | 0.72, 1.21 | 0.5 | 1.18 | 0.49, 3.05 | 0.7 |
| Bareground | 12,341 | 1.03 | 0.71, 1.32 | 0.8 | 1.49 | 0.79, 2.47 | 0.2 |
| Rangeland | 12,341 | 0.99 | 0.76, 1.22 | >0.9 | 1.22 | 0.58, 2.37 | 0.6 |
| LST_10 | 12,341 | 0.56 | 0.50, 0.63 | <0.001 | 0.21 | 0.06, 0.69 | 0.010 |
| LST_11 | 12,341 | 0.60 | 0.53, 0.67 | <0.001 | 3.34 | 1.02, 11.0 | 0.046 |
| EVI | 12,341 | 2.06 | 1.85, 2.30 | <0.001 | 3.93 | 0.72, 21.6 | 0.11 |
| NDVI | 12,341 | 1.98 | 1.78, 2.19 | <0.001 | 0.51 | 0.08, 3.33 | 0.5 |
| TCB | 12,341 | 1.01 | 0.88, 1.16 | >0.9 | 0.41 | 0.23, 0.71 | 0.002 |
| TCG | 12,341 | 1.88 | 1.70, 2.07 | <0.001 | 0.97 | 0.30, 3.07 | >0.9 |
| TCW | 12,341 | 1.23 | 1.06, 1.44 | 0.008 | 0.49 | 0.26, 0.88 | 0.019 |
| Built_c | 12,341 | 0.83 | 0.78, 0.89 | <0.001 | 1.11 | 0.98, 1.26 | 0.11 |
| Built_h | 12,341 | 0.67 | 0.62, 0.73 | <0.001 | 1,070,681 | 8.85, 197,874,120,850,197 | 0.078 |
| Built_s | 12,341 | 0.65 | 0.59, 0.70 | <0.001 | 0.84 | 0.57, 1.23 | 0.4 |
| Built_v | 12,341 | 0.67 | 0.62, 0.72 | <0.001 | 0.00 | 0.00, 0.16 | 0.086 |
| Pop | 12,341 | 0.76 | 0.72, 0.81 | <0.001 | 0.83 | 0.74, 0.93 | 0.001 |
| ^1^OR = Odds Ratio, CI = Confidence Interval | | | | | | | |
