## Supplementary figure 3 for "Fine scale spatial mapping of urban malaria prevalence for microstratification in an urban area of Ghana"

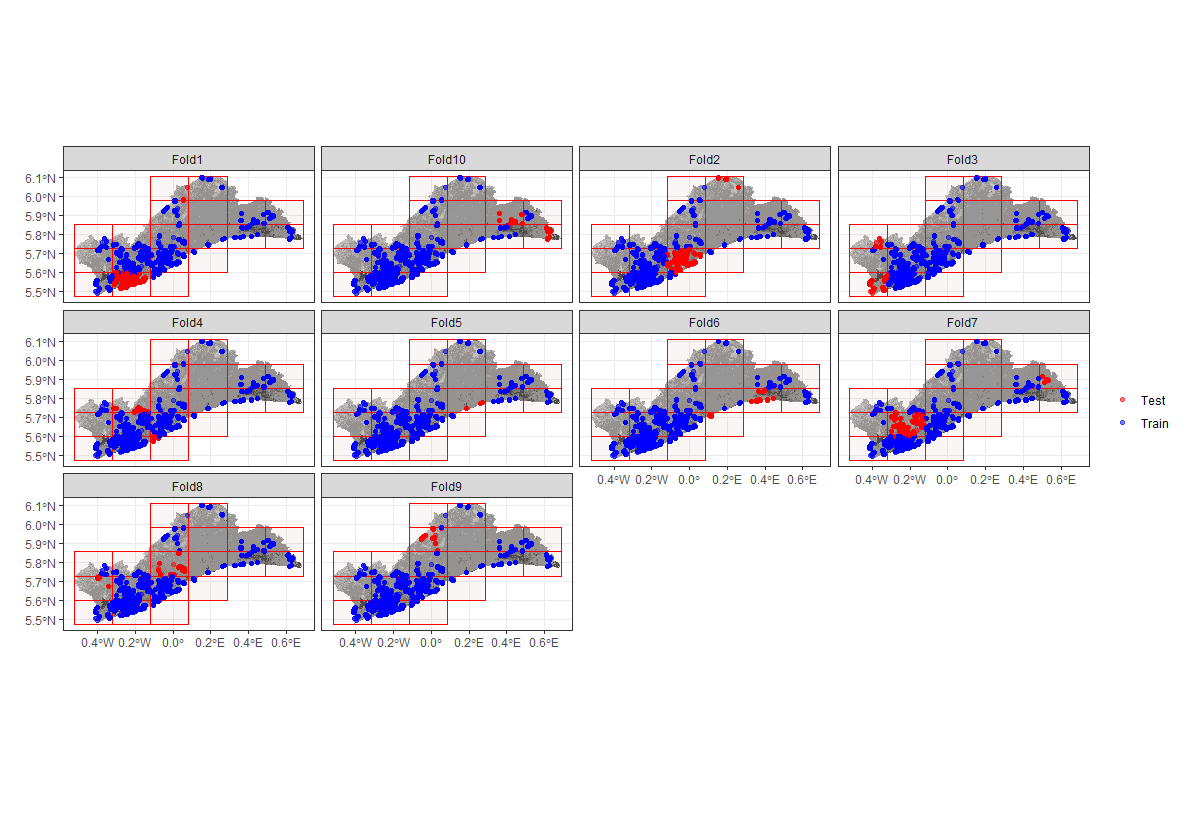

Supplementary figure 3: Spatial assignment of data into 10-folds for cross validation. Red dots denote samples assigned as test set and blue dots denote samples assigned as train set.
