## Supplementary table 3 for "Fine scale spatial mapping of urban malaria prevalence for microstratification in an urban area of Ghana"

Supplementary table 3: Number of observations for each fold for random and spatial blocking cross-validations

| Fold ID | Random cross validation | Spatial blocking cross validation | |
| --- | --- | --- | --- |
|  |  | Test set | Train set |
| 1 | 1238 | 2456 | 9915 |
| 2 | 1237 | 376 | 11995 |
| 3 | 1237 | 2399 | 9972 |
| 4 | 1237 | 203 | 12168 |
| 5 | 1237 | 349 | 12022 |
| 6 | 1237 | 533 | 11838 |
| 7 | 1237 | 1005 | 11366 |
| 8 | 1237 | 4024 | 8347 |
| 9 | 1237 | 438 | 11933 |
| 10 | 1237 | 588 | 11783 |
