## Supplementary table 2 for "Fine scale spatial mapping of urban malaria prevalence for microstratification in an urban area of Ghana"

Supplementary table 2: Final set of covariates and their VIF values

| Covariates | VIF value |
| --- | --- |
| Water | 1.059 |
| Trees | 1.177 |
| Built_area | 5.427 |
| Bareground | 1.537 |
| Rangeland | 4.825 |
| LST_11 | 1.664 |
| EVI | 2.340 |
| TCB | 2.970 |
| TCW | 3.395 |
| PC1 | 2.700 |
| PC2 | 1.234 |
| PC3 | 1.459 |
| PC4 | 1.139 |
