## Supplementary figure 2 for "Fine scale spatial mapping of urban malaria prevalence for microstratification in an urban area of Ghana"

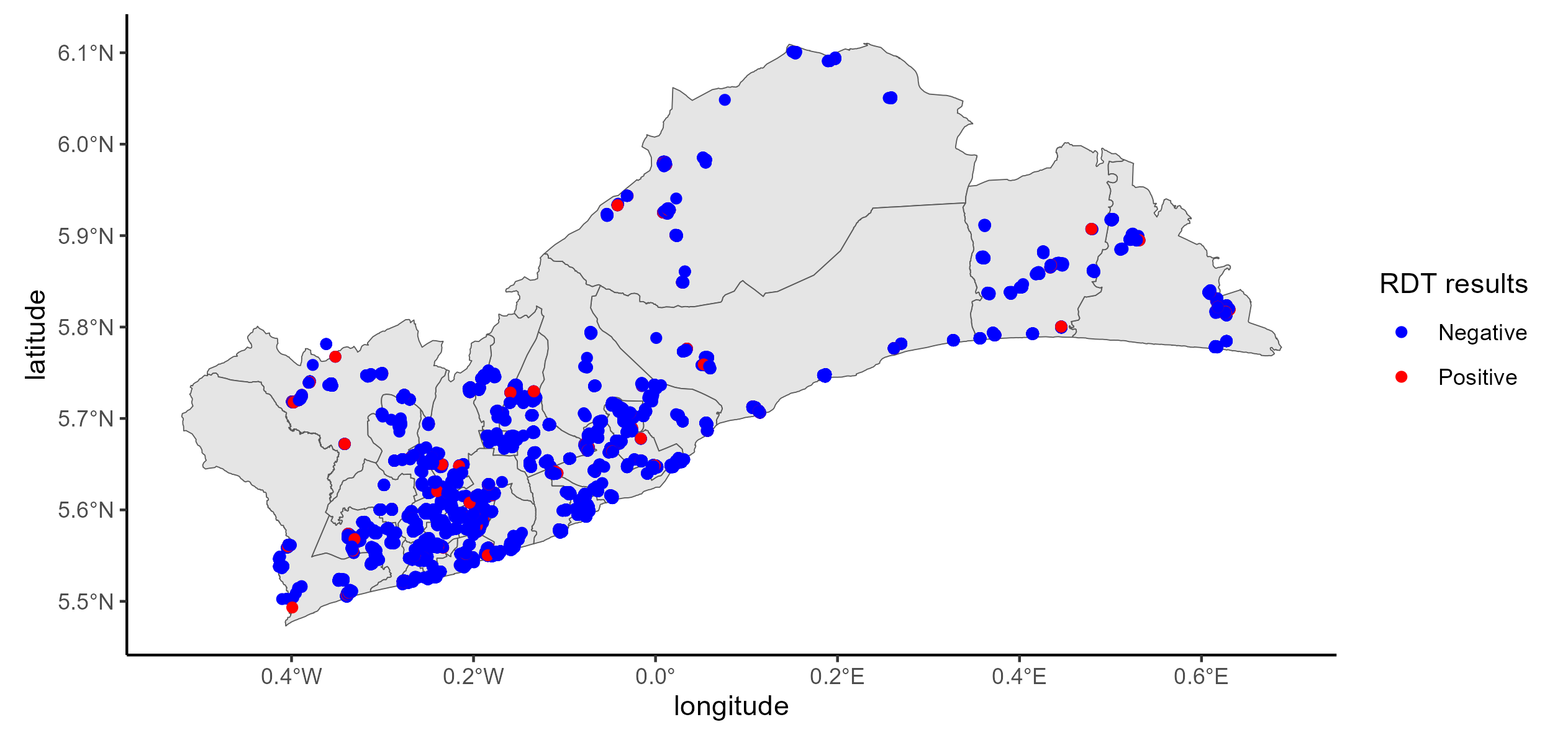

Supplementary figure 2: Distribution of RDT results of study participants. Blue dots indicate negative mRDT results while red dots indicate positive resul.t
