## Supplementary table 1 for "Fine scale spatial mapping of urban malaria prevalence for microstratification in an urban area of Ghana"

Supplementary table 1: Description of covariates, source, and spatial resolution

| Covariate | Description | Sources | Spatial resolution |
| --- | --- | --- | --- |
| Land cover classes |  |  |  |
| Water | Water | LANDSAT/LC08/C02/T1 | 30m |
| Trees | Trees | LANDSAT/LC08/C02/T1 | 30m |
| Crops | Crops | LANDSAT/LC08/C02/T1 | 30m |
| Built-area | Built area | LANDSAT/LC08/C02/T1 | 30m |
| Flooded_veg | Flooded vegetation | LANDSAT/LC08/C02/T1 | 30m |
| Bareground | Bare ground | LANDSAT/LC08/C02/T1 | 30m |
| Rangeland | Rangeland | LANDSAT/LC08/C02/T1 | 30m |
| Environmental/ climatic |  |  |  |
| EVI | Enhanced vegetation index | MODIS derivative(25) | 30m |
| NDVI | Normalized difference vegetation index | Landsat 8-USGS | 30m |
| LSTemp_10 | Land surface temperature band 10 | MODIS derivative | 30m |
| LSTemp_11 | Land surface temperature band 11 | MODIS derivative | 30m |
| TCB | Tasselled cap brightness; measure of reflectance | MODIS derivative | 30m |
| TCG | Tasselled cap greenness | MODIS derivative | 30m |
| TCW | Tasselled cap wetness | MODIS derivative | 30m |
|  | Urban covariates |  |  |
| GHS-BUILT-H | GHS built-up height | Global Human settlement layer (GHSL) | 100m |
| GHS-BUILT-S | GHS built-up surface | Global Human settlement layer (GHSL) | 100m |
| GHS-BUILT-V | GHS built-up volume | Global Human settlement layer (GHSL) | 100m |
| GHS-BUILT-C | GHS built-up characteristics | Global Human settlement layer (GHSL) | 100m |
| GHS-POP | GHS population grid (R2023) | Global Human settlement layer (GHSL) | 100m |
